# Attacks on Ukrainian healthcare facilities during the first year of the full-scale Russian invasion of Ukraine

**DOI:** 10.1101/2023.08.29.23294763

**Authors:** Dennis G. Barten, Derrick Tin, Fredrik Granholm, Diana Rusnak, Frits van Osch, Gregory Ciottone

## Abstract

**Background:** Although the Geneva Conventions and Rome Statute demand protections for healthcare facilities during war, breaches of these protections are frequently reported. The ongoing war in Ukraine is no exception, with several healthcare attacks eliciting widespread condemnation. The Ukrainian Healthcare Center (UHC) has been collecting, verifying and documenting attacks on health infrastructure since the Russia-Ukraine War was launched. The aim of this study was to assess UHC documented healthcare facility attacks during the first year (24 February 2022 to 25 February 2023) of the full-scale Russian invasion of Ukraine.

**Methods:** The Berkeley Protocol on Digital Open Source Investigations was used to document healthcare attacks. Data collection included temporal factors, location, facility type, attack and weapon type, number of killed and injured healthcare personnel and civilians, and whether facilities were damaged, destroyed or attacked more than once.

**Results:** There were 334 documented attacks on 267 Ukrainian healthcare facilities, with 230 facilities being damaged and 37 destroyed. General hospitals, primary care clinics, emergency departments and children’s hospitals were most frequently targeted. The majority of attacks took place during the first three months and in eastern Ukrainian oblasts. Heavy weaponry was employed in almost all attacks. The total number of casualties included 97 fatalities and 114 injuries.

**Conclusions:** During the first year of the full-scale Russian invasion of Ukraine, there were 334 attacks on 267 Ukrainian healthcare facilities documented by the UHC. Heavy weaponry was commonly used, and the direct impact of attacks was considerable in terms of facility damage and casualty tolls.

## Introduction

The Russia-Ukraine War has been ongoing since February 2014, when Russia annexed the Crimean Peninsula from Ukraine and supported pro-Russian separatists in the Donbas region. Throughout 2021, tensions rose due to Russian military buildup near the border with Ukraine. On 24 February 2022, the war significantly escalated as Russia launched a full-scale invasion of Ukraine, attracting worldwide media attention and eliciting widespread condemnation. Over the course of the full-scale invasion, several territories in the northeast, east and southeast of Ukraine have been temporarily occupied.^1^

Although the Geneva Conventions and Rome Statute demand protections for healthcare workers and healthcare facilities during war, breaches of these protections have been reported during multiple armed conflicts in recent years, including the Chechen War (1999-2009), Syria Civil War (2011-present) and Tigray War in Ethiopia (November 2020-present).^2-5^ The World Health Organization (WHO) stated in 2022 that the targeting of healthcare facilities now has become part of the strategy and tactics of warfare in conflict zones,^6^ and condemns such acts as a violation of international humanitarian law (IHL).^7^ Attacks on healthcare are clearly being observed during the ongoing war in Ukraine, raising significant concerns amongst health experts globally.^8,9^

Beyond the direct effects of physical and psychological injuries, deaths, and destruction of healthcare infrastructure, attacks on healthcare lead to wider disruptions of routine and acute emergency care, maternal and child health, and may enhance the spread of infectious diseases, including SARS-CoV-2, hepatitis, tuberculosis, and human immunodeficiency virus (HIV).^10-12^ The prevention of attacks on healthcare facilities is of paramount importance and documentation is essential to identify violations, create mechanisms for protection and accountability, and develop the political will to enforce them.^13,14^ The Ukrainian Healthcare Center (UHC), a think tank that played a significant role in the health system reform in Ukraine in 2016-2019, concentrated on war-related initiatives after the 2022 Russian invasion of Ukraine, including collecting, verifying and documenting attacks on healthcare infrastructure.^15^

The aim of this study is to assess all healthcare facility attacks that were documented by the UHC during the first year (24 February 2022 to 25 February 2023) of the full-scale Russia-Ukraine War. This information can be used to identify patterns of healthcare attacks in order to develop strategies to avoid or mitigate these disruptions to healthcare delivery, and better understand the impact of war on a country’s health system.

## Methods

The *Ukrainian Healthcare Center* is a think tank based in Kyiv, Ukraine, providing consultancy, analytics, and educational services.^15^ Its core competencies include health system policy and governance, health economics and financing, and health system transformation. UHC started the documentation of attacks on healthcare infrastructure in February 2022, following the onset of the full-scale Russian invasion.

### Data collection

The Berkeley Protocol on Digital Open Source Investigations was used in this study. The protocol identifies international standards for conducting online research of alleged violations of international criminal, human rights, and humanitarian law, with representatives of the United Nations engaged in the documentation process.^16^ The protocol provides guidance on methodologies and procedures for gathering, analyzing, and preserving digital information in a professional, legal, and ethical manner. Information on attacks on healthcare facilities in Ukraine between 24th February 2022-25th February 2023 were collected through open sources such as media outlets, social media platforms (Facebook, Twitter, Instagram, Telegram and YouTube), official government websites, witnesses and physical, in-person site visits. To ensure thoroughness, additional cross-checking was conducted with existing databases on attacks and damage to healthcare facilities to identify potential overlooked cases. These databases were maintained by the Ministry of Health of Ukraine (MoH), Insecurity Insight, and Médecins Sans Frontières (MSF). The collected information, where available, included: facility name; facility type; official address and geographical coordinates; photo evidence from open sources and/or witnesses; testimonies from witnesses (audio, chats, SMS); photo or audio recordings from site visits; satellite image analysis; and the reported number of killed and injured healthcare personnel. It was also recorded if healthcare facilities were attacked more than once. For the purpose of this study, the database was last accessed on May 8, 2023.

### Definitions

Healthcare facilities were defined as core health system institutions staffed with doctors and/or nurses. Rural feldsher or midwife points, pharmacies, and recreational facilities were not included. An attack on a healthcare facility was a priori defined as any form of physical violence or obstruction that interferes with the accessibility and delivery of healthcare by these facilities. A facility was considered damaged if it was still partially functioning and a facility was considered destroyed if it was completely non-functioning. Weapon and attack types were determined by the definitions used in the Explosive Weapons Monitor.^17^ Ground launched explosive weapons are launched from any surface-level platform, including weapons thrown by a person, or fired from warships or vehicles. These include artillery shells (projectiles fired from a gun, cannon, howitzer, or recoilless rifle), tank shells, ground-launched missiles, mortars, rockets (typically missiles which do not contain guidance systems), non-specific shelling, rocket-propelled grenades, and hand grenades. Air-launched explosive weapons include any weapon fired from a rotary or fixed-wing aircraft, including unmanned aerial vehicles or drones. These include air-dropped bombs (bombs reported as being delivered by air), airstrikes (attacks from a helicopter, drone, or plane), and missiles or rockets launched from an aircraft. Directly-emplaced explosive weapons include any that are brought to the facility and detonated. Such weapons consist of improvised explosive devices of all types, including vehicle-borne improvised devices, landmines and explosive vests. For each attack the facility type was determined. ‘Other facility types’ included dental clinics, monoprofile (specialty) hospitals, addiction or rehabilitation centers and nursing homes.

### Verification

The system of verification for the database on healthcare facility attacks compromised two levels contingent upon the quantity of evidence collected. There are four types of verifying evidence: 1. News reporting about the attack (including social media posts); 2. Graphic evidence (photo or video of damage / destruction); 3. Witness testimony (including information collected via phone calls); 4. Satellite imagery (the imagery or analysis that stated the facility was damaged). If one type of evidence was obtained, level 1 verification was reached. Two or more types of evidence resulted in verification level 2.

### Data analysis

All information was collected into a Google Spreadsheet (Google Inc., Mountain View, California, USA), and sorted and analyzed based on the date of the event, location, facility type, weapon type used, medical staff and civilian injury and death tolls, and whether the facility was damaged, destroyed or attacked more than once.

Ethical approval of this study was waived by the medical-ethical review board of Maastricht University Medical Center based on Dutch legislation for research concerning humans (Maastricht, The Netherlands; 2022-3518).

## Results

### Attacks on healthcare facilities

There were 334 documented attacks on 267 Ukrainian healthcare facilities between 24 February 2022 and 25 February 2023. A verification level of 2 was reached in 62.3% of the attacks. Thirty-eight facilities (38/267 facilities; 14.2%) were attacked more than once (total: 105 attacks on 38 facilities; median 2 attacks; range 2-8 attacks). The most frequently targeted facility types were general hospitals (114/334 attacks; 34.1%), followed by primary care clinics (65 attacks, 19.5%), other facility types (37; 11.1%), emergency departments (23; 6.9%), children’s hospitals (20; 6.0%), maternity hospitals (20; 6.0%) and outpatient facilities (17; 5.1%). Attacks stratified by facility type are presented in Figure 1.

**Figure 1:**
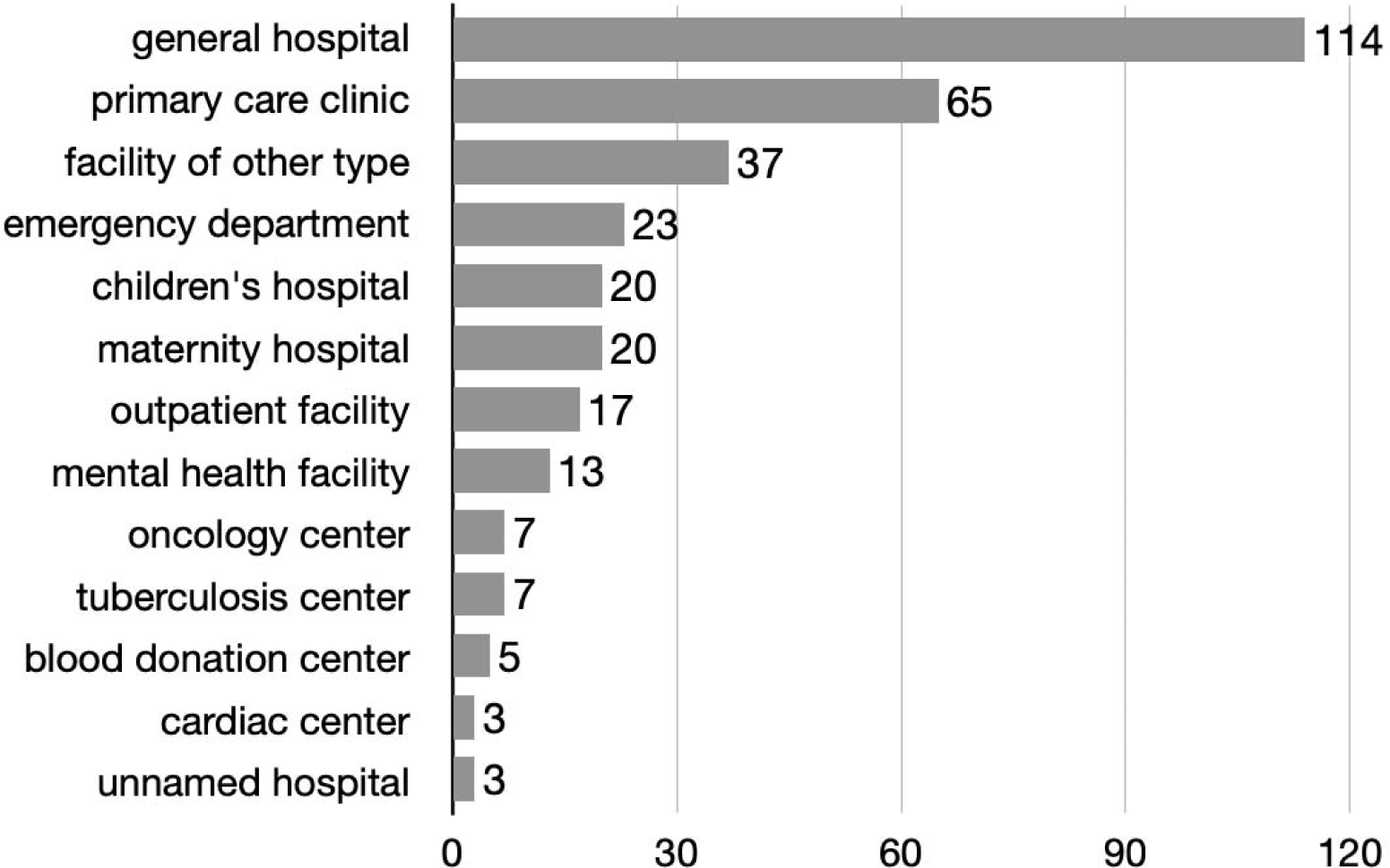
Attacks stratified by facility type.

### Weapon and attack types

Explosives were the most frequently documented weapon type (241 attacks; 72.1%). In 2 attacks firearms were used (2; 0.6%) and a combination of explosives and firearms was used in 1 attack (0.3%). There was no information about the weapon type used in 90 attacks (27%). Attack types mostly included ground launched attacks (93 attacks; 27.8%) or air launched attacks (45 airstrikes; 13.5%). There was one attack with a directly-emplaced explosive and a combination of attacks was reported in 5 attacks (1.5%). There was no verified information on attack types in 190 attacks (56.9%). The total of 334 attacks resulted in 230 facilities (86.1%) being damaged and 37 facilities (13.9%) destroyed.

### Injuries and deaths

In total, 9 healthcare workers and 105 civilians were killed in the attacks, with a further 26 healthcare workers and 88 civilians injured. The single attack with the highest number of casualties (56 deaths) was the Stara Krasnianka care house attack on 11 March 2022 near Kreminna, Luhansk Oblast. Another large-scale attack was the Mariupol maternity hospital airstrike on 9 March 2022. This attack resulted in 6 deaths and 33 people with injuries.

### Temporal distribution of attacks

Most attacks occurred during the early phase of the full-scale Russian invasion of Ukraine, with 22 (6.6%), 163 (48.8%) and 36 (10.8%) incidents in February, March and April 2022, respectively. The temporal distribution of attacks per month is shown in Figure 2. The number of facility attacks per month during the total study period equaled 27.8.

**Figure 2:**
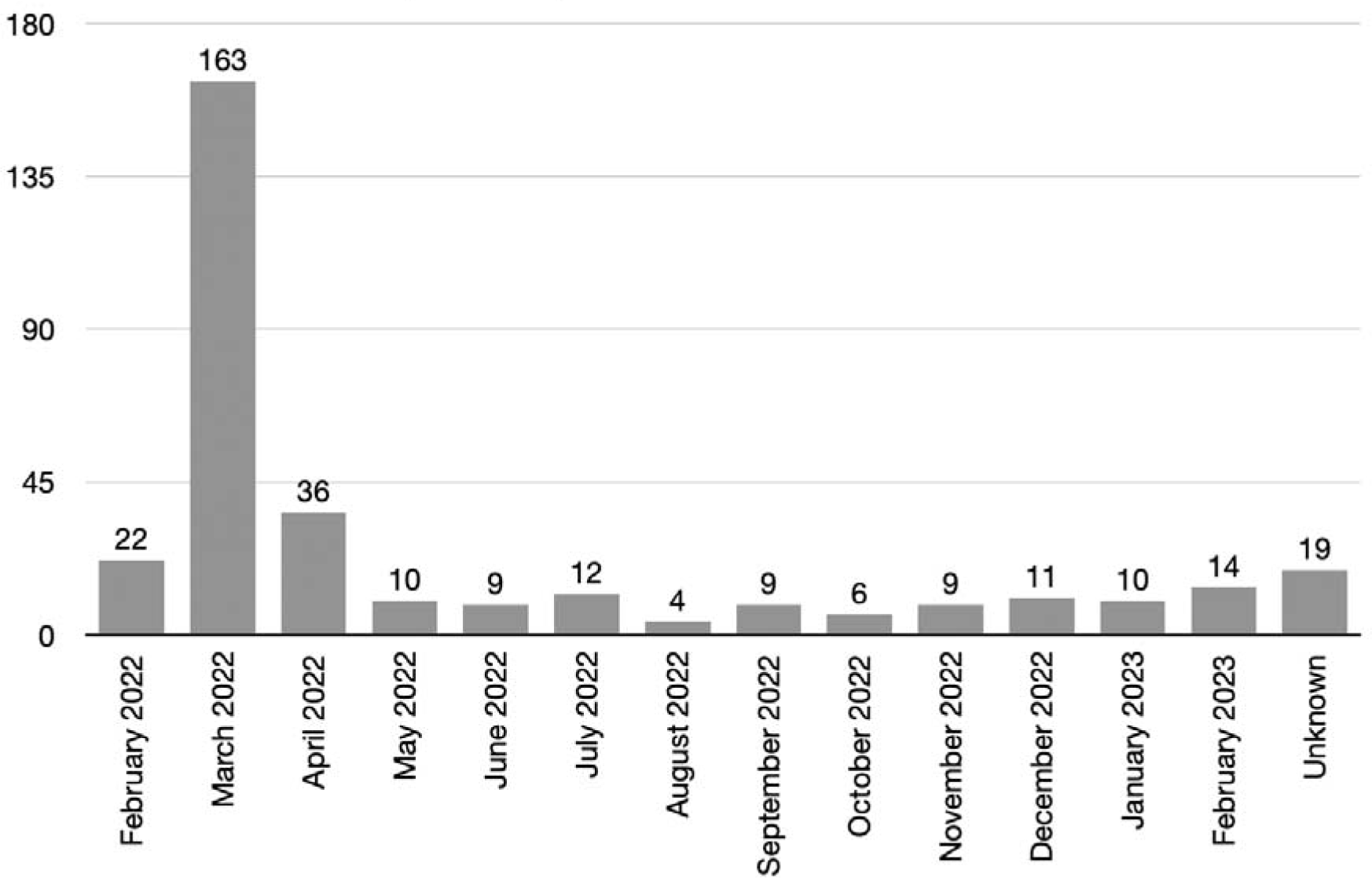
Temporal distribution of healthcare facility attacks, per month.

### Regional distribution of attacks

Attacks occurred in 13 out of 24 Ukrainian Oblasts. Most attacks were reported in Kharkiv Oblast (85 attacks, 25.5%), followed by Donetsk Oblast (61; 18.3%), Kyiv Oblast (47; 14.1%), Luhansk Oblast (31; 9,3%) and Kherson Oblast (29; 8.7%). The geographic distribution of attacks is visualized in Figure 3, and the number of attacks per oblast is depicted in Figure 4.

**Figure 3:**
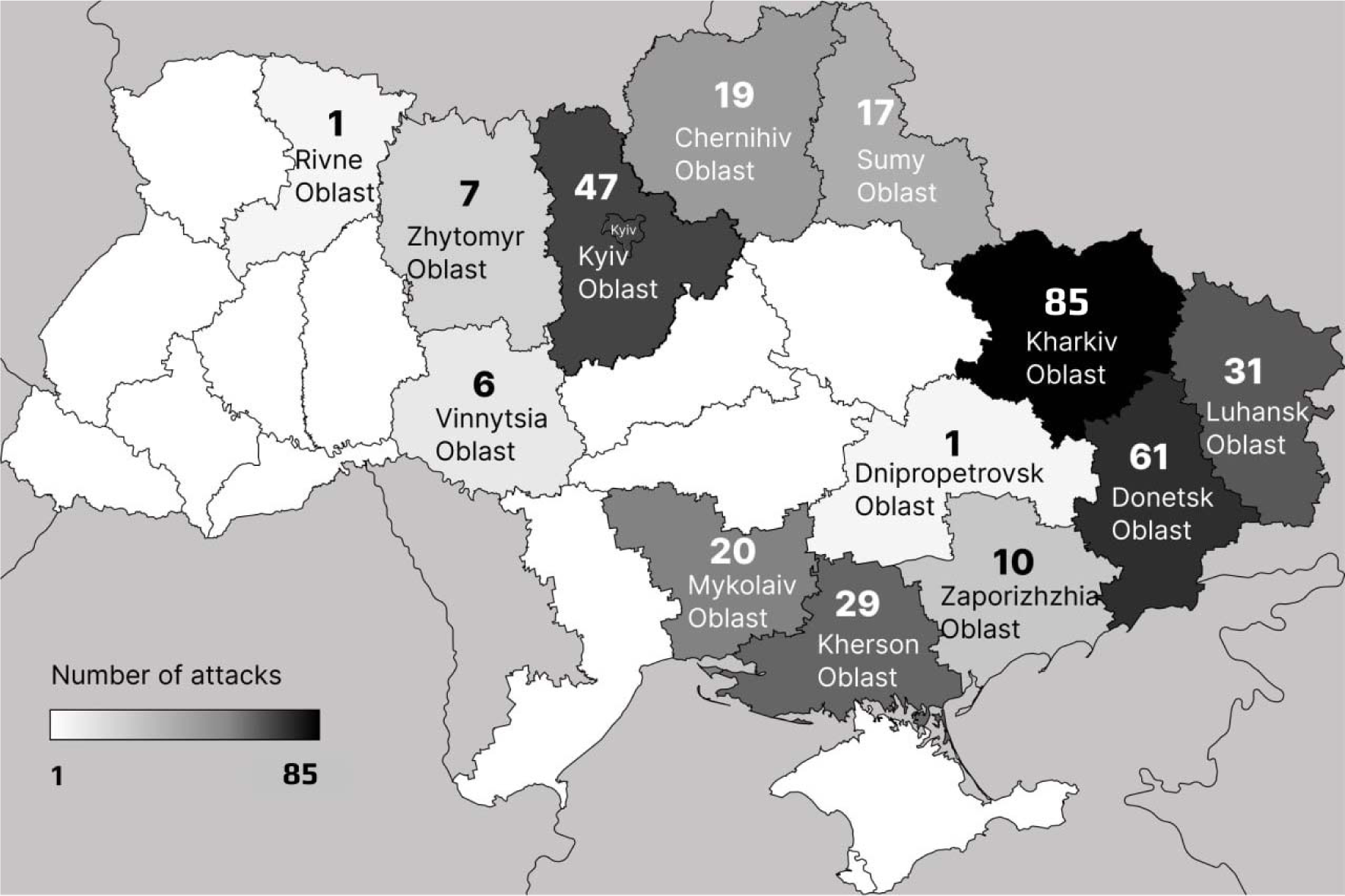
Geographic distribution of attacks on healthcare facilities in Ukraine.

**Figure 4:**
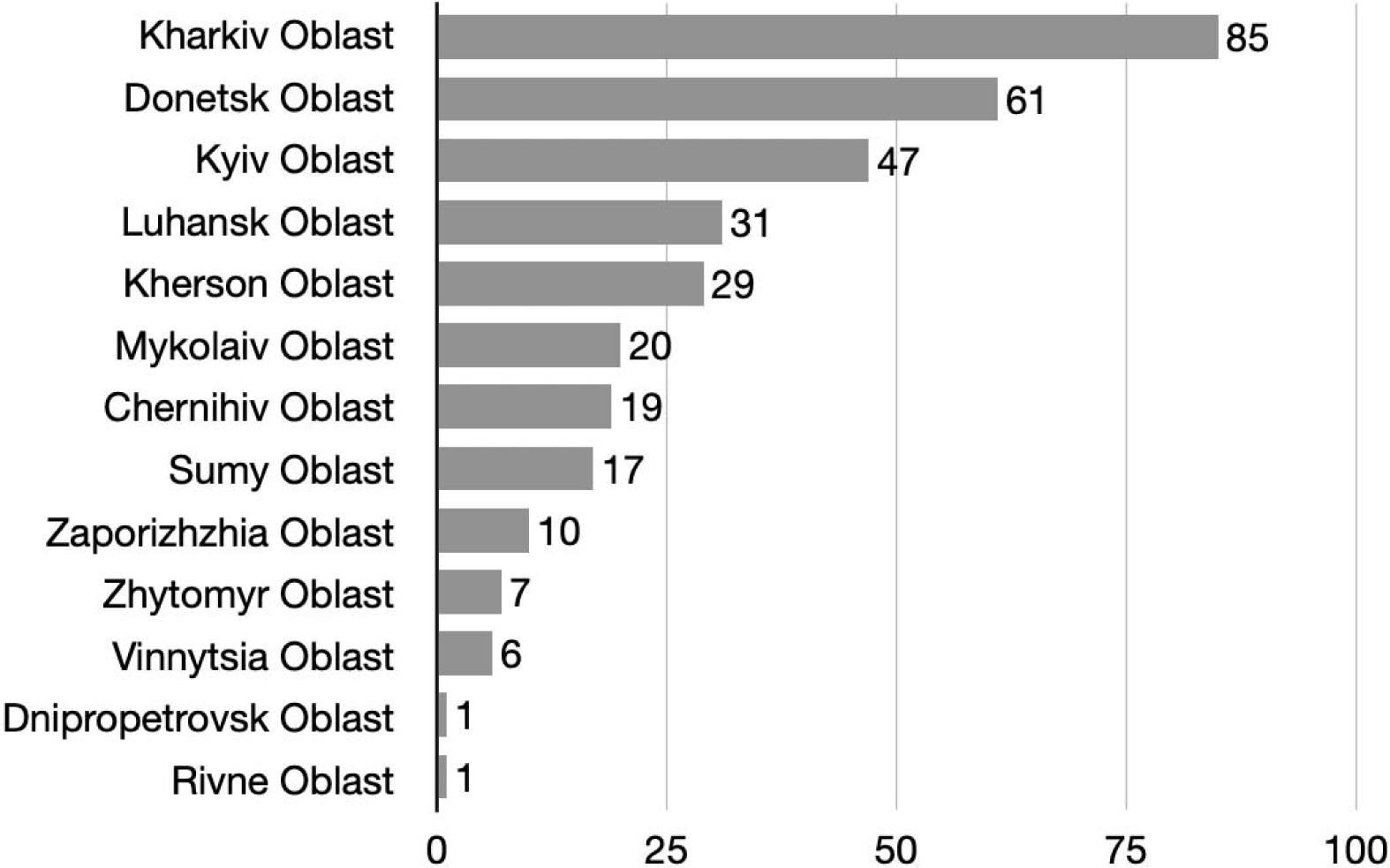
Distribution of healthcare facility attacks per oblast.

## Discussion

Attacks on healthcare in any setting are deadly and disruptive. When they occur during a conflict or war, they also dramatically limit the capability and capacity of the healthcare system to provide care for future casualties. During the first year of the full-scale Russian invasion of Ukraine, there were 334 attacks on 267 Ukrainian health facilities documented by the UHC. The majority of attacks were targeted at general hospitals and primary care clinics. The attacks were predominantly concentrated in the eastern oblasts of Ukraine, occurring mainly in the initial months following the full-scale invasion. Heavy weaponry was commonly used, and the direct impact of attacks was considerable in terms of facility damage and casualty tolls. Although healthcare attacks were condemned worldwide early in the conflict, they continued to be perpetrated throughout the entire study period.

The 2022 full-scale Russian invasion of Ukraine was not the only phase of the ongoing war in which healthcare attacks were observed. Researchers previously determined that over one-third of the hospitals and clinics in the Donbas region had been damaged or destroyed from 2014 through 2017. The most significant damage occurred in areas where the fighting was most intense – that is, along the so-called “line of contact” dividing Ukrainian government-held territory and that occupied by Russia-backed separatists.^18^ During the first year of the full-scale invasion, several governmental and non-governmental organizations have been documenting attacks on Ukrainian healthcare facilities, including the MoH of Ukraine, World Health Organization (WHO), MoH, Physicians for Human Rights, Insecurity Insight and MSF.. The number of reported attacks varies considerably between databases. For example, the WHO reported 1,033 attacks during the first year, the MoH claimed that more than 1,200 medical facilities were damaged or destroyed, and a joint report of five non-governmental organizations, based both inside and outside Ukraine, included 707 attacks on Ukraine’s healthcare system.^19-21^ These differences can be explained by a different handling of several definitions, and whether incident verification was applied. For example, what is considered a healthcare facility; when is there enough damage to be included; and what is considered an attack? The UHC method is characterized by the use of strict definitions and incident verification. All in all, it is undisputed that Ukraine witnessed an unprecedented scale of healthcare attacks. It is estimated that one in 10 of Ukraine’s hospitals have been directly damaged from attacks, and in cities such as Mariupol nearly all health facilities were harmed in some way.^19,22^ Furthermore, dozens of hospitals were attacked multiple times, underscoring not only the indiscriminate nature of attacks but also the possibility that they were deliberately targeted.^14,19^

Attacks on healthcare during armed conflicts are relatively understudied. For some recent conflicts attempts were made to estimate the impact of healthcare attacks. These estimates vary from 0.12 healthcare facility attacks per month in Iraq to 4.65, 6.67 and 9.64 attacks per month in Yemen, Kosovo and Syria, respectively.^4^ In contrast, the number of attacks on Ukrainian healthcare facilities per month equaled a staggering 27.8.

Unfortunately, attacks on healthcare are common events in recent armed conflicts and hybrid wars. Disrupting the healthcare system in a country using hybrid attacks has the ultimate aim of destabilizing trust in government and key organizations, but also directly reduces the effectiveness and capabilities of healthcare as a key strategic resource.^8,23^ In this aspect, prehospital care and emergency medicine are main targets of aggression, since a decrease in their capacity would seriously affect a country’s ability to care for war casualties.^23^

Ensuring the protection of healthcare, encompassing access to and delivery of healthcare services, is a joint responsibility that falls upon all parties engaged in an armed conflict. States, armed groups, and other involved parties bear a shared duty to uphold IHL in this regard. Beyond protection there is a pressing need to ensure accountability, which helps deter war crimes, promote justice for victims, and prevent future violations. Despite the adoption of United Nations Security Council Resolution 2286 in 2016, which condemns attacks against health facilities and personnel and calls for prevention and accountability, healthcare attacks persist.^2,5^ In order to uphold these rules and regulations, it is essential to strengthen international mechanisms for accountability, promote improved investigational platforms and methodologies, support domestic efforts to hold perpetrators accountable, while also increasing awareness and advocacy for the protection of healthcare in times of war and conflict.^19,24-26^

### Limitations

The UHC database solely records attacks against essential healthcare facilities where medical professionals (doctors and nurses) provide aid. It does not include attacks on public health institutions, paramedics and midwifery facilities, drug warehouses, sanatoriums, pharmacies, or other medical-related establishments. This criterion may result in a lower count of attacked facilities compared to other databases that document all healthcare-related attacks. Furthermore, indirect attacks on healthcare, such as critical infrastructure attacks, were not listed in this database. Finally, attacks on ambulances were not included in this analysis mainly due to the challenge of verifying the data. However, it is important to acknowledge that this exclusion could be seen as a limitation, as attacks on ambulances can significantly disrupt healthcare services.^27,28^

The database remains dynamic, continuously updated with new information on attacks as it emerges. Consequently, the number of attacks and their details can change over time. For the purpose of this study, the last updated version was May 8, 2023, and since then, certain details have been revised or updated, but these changes are not reflected in the analysis presented in this study.

The UHC database relies on verifiable evidence. Consequently, the figures presented in this study offer a conservative estimate of healthcare attacks in Ukraine and are likely to be an underestimation. Similar to any other incident data collection, this dataset is subject to selective reporting. Various factors, such as limited access, inadequate information, unavailable internet connections, and other omissions, may lead to the inclusion or exclusion of certain events. Notably, incidents that took place in territories directly or previously occupied by Russia were infrequently or rarely reported, leading to a partial portrayal of the extent of violence in these specific regions.

During on-site visits, significant and crucial information about the attacks was gathered. However, it is important to note that most of these visits occurred a month or more after the territories were liberated, which also corresponds to the time of the attacks. This timing could potentially affect the amount of evidence collected on-site and the estimation of damage, as some facilities might have been partially repaired by the time of the visit.

. Although there are several limitations to the database used for this study, the database is probably the most comprehensive dataset of on healthcare facilities in this war. While summary estimates of healthcare damage within individual conflict zones are typically readily available, many lack a carefully delineated methodology, precise periodicity, and detailed information on location, yielding wide variations in estimates.^25^

## Conclusion

During the first year of the full-scale Russian invasion of Ukraine, there were 334 attacks on 267 Ukrainian health facilities documented by the UHC. Heavy weaponry was commonly used, and the direct impact of attacks was considerable in terms of facility damage and casualty tolls. Despite global condemnation of these healthcare attacks early in the war, they continued to be perpetrated.

## Data Availability

All data produced in the present study are available upon reasonable request to the authors

